# Classification of Functional Movement Disorders with Resting State fMRI

**DOI:** 10.1101/2021.09.01.21262677

**Authors:** Rebecca E. Waugh, Jacob A. Parker, Mark Hallett, Silvina G. Horovitz

## Abstract

Functional movement disorder (FMD) is a type of functional neurological disorder characterized by abnormal movements that patients do not recognize as self-generated. Prior imaging studies show a complex pattern of altered activity, linking regions of the brain involved in emotional responses, motor control, and agency. This study aimed to better characterize these relationships by building a classifier via support vector machine (SVM) to accurately classify 61 FMD patients from 59 healthy controls using features derived from resting state functional MRI (rs-fMRI). First, we selected 66 seed regions based on prior related studies, then calculated the full correlation matrix between them, before performing recursive feature elimination to winnow the feature set to the most predictive features and building the classifier. We identified 29 features of interest that were highly predictive of FMD condition, classifying patients from controls with 80% accuracy. The features selected by the model highlight the importance of the interconnected relationship between areas associated with emotion, reward and sensorimotor integration, potentially mediating relationships between regions associated with motor function, attention and executive function. Exploratory machine learning was able to identify this distinctive, abnormal pattern, suggesting that alterations in functional linkages between these regions may be a consistent feature of the condition in many FMD patients.

## Introduction

Functional movement disorder (FMD) is characterized by abnormal movement and is a sub-type of functional neurological disorder (FND). FMD patients perceive these movements as involuntary, and this lack of self-agency discriminates the disorder from feigning or malingering (Czarnecki & Hallett, 2012). Functional neuroimaging studies indicate that patients with FMD have abnormal brain function. Neural activity associated with FMD symptomology differs from that of feigned symptoms (Cojan et al., 2009; Spence et al., 2000; Stone et al., 2007; van Beilen et al., 2011; Voon, Gallea, et al., 2010) or organic dystonia (Schrag et al., 2013). Despite heterogeneity in study design and criteria for subject inclusion, altered activity in multiple brain regions has been consistently implicated in FMD, including primary motor, supplementary motor area (SMA), prefrontal cortex (PFC), amygdala, and temporo-parietal (TPJ), and parietal-occipital (POJ) junctions (Voon et al., 2016).

When compared to healthy controls, FMD cohorts tend to show reduced activity in pre-motor, supplementary motor, and primary motor cortices during motor execution (Dogonowski et al., 2019; Marshall et al., 1997; Voon et al., 2011) and movement observation tasks (Burgmer et al., 2006). A lack of habituation to and stronger arousal in limbic areas, particularly the amygdala, in response to emotional stimuli is also present in FMD patients (Aybek et al., 2015; Hassa et al., 2017; Voon, Brezing, et al., 2010), along with atypical activity in parts of inferior frontal, temporal, and occipital regions (Espay et al., 2018). Altered connectivity has been detected between areas associated with motor planning, attention, and self-monitoring, such as the temporo-parietal-occipital junctions, prefrontal cortices, and motor regions (Diez et al., 2019; Maurer et al., 2016).

It has been postulated that overactivation in areas associated with emotional processing suppresses and interferes with motor execution (Hassa et al., 2017; Kanaan et al., 2007; Voon et al., 2011), resulting in a discrepancy between motor planning and execution that impacts the FMD patient’s sense of agency over his or her own actions (Roelofs et al., 2019). Altered connectivity in the right TPJ, an area associated with feelings of self-agency (Nahab et al., 2011; Nahab et al., 2017), has been strongly implicated in functional imaging studies of FMD patients, suggesting it may be an intrinsic feature of the condition (Aybek et al., 2014; Baek et al., 2017; Maurer et al., 2016). Other areas associated with volitional motor control are also impacted, including the pre-SMA and dorsolateral PFC (dlPFC) – supporting the FMD patient’s perception of the involuntariness of his or her symptoms (Spagnolo et al., 2021). This abnormal interplay between networks associated with emotion, motor planning and action, and attention, self-monitoring, and agency characterizes FMD (Monsa et al., 2018). Successful treatment of FMD is associated with the disruption of this pattern (Dogonowski et al., 2019; Faul et al., 2020).

Although it is evident that FMD patients have abnormal neural activity across multiple regions and brain networks, patients are also a heterogenous group in terms of symptoms and frequently present with comorbid neurological or psychiatric disorders. Diagnosis of FMD should be made based on positive symptoms, such as entrainment or Hoover’s sign, that are not consistent with another neurological disease (American Psychiatric Association, 2013). However, although FMD patients seeking care at neurology clinics are common, reaching an appropriate diagnosis can be difficult. It can take two to ten years following symptom onset to make the diagnosis (Perez et al., 2021), and most neurologists continue to order additional testing to make a ‘diagnosis of exclusion’ (LaFaver et al., 2020).

Wegrzyk and colleagues proposed that machine learning classification might be helpful in making diagnoses and developing biomarkers for FND. They achieved 68% classification accuracy by training a support vector machine (SVM) classification algorithm with measures derived from rs-fMRI in 26 FND patients and 27 healthy controls, indicating that rs-fMRI contains relevant information about the disorder that can be detected in a supervised learning paradigm. Increased connectivity between the right caudate and left amygdala and postcentral gyri, along with decreased connectivity between the right TPJ and frontal regions characterized the most predictive features for the model (Wegrzyk et al., 2018).

The most common use of machine learning classifiers in a clinical setting is to learn to predict the group assignment of subjects from their quantitative measurements. While this is done by considering all the measurements that differ systematically between groups, they can also indicate which measurements in a particular subject led to a correct prediction. As such, these techniques have the potential to be useful in FMD to improve diagnostic clarity. Additionally, such models are sometimes more able than other techniques to detect complex and subtle relationships between features. Here, we were interested in using a machine learning approach with a dataset of 120 samples consisting of FMD patients and healthy controls.

The goals of this study were two-fold. First, we aimed to build a highly accurate classifier based on rs-fMRI data to distinguish between FMD patients and healthy controls. Second, by investigating which features were most important to the model, we explored which patterns of activity are more relevant and consistently predictive in a broad range of FMD patients. As such, the initial feature list was highly inclusive, derived from a range of prior fMRI studies in FMD patients, including resting state and task-based imaging paradigms. Then, the full feature list underwent a feature selection process to produce a more limited set of features that produced the highest predictive value to the classifier.

## Methods and Materials

### 2.1 Participants

61 patients with clinically definite FMD recruited by the Human Motor Control clinic and 59 age and gender matched healthy controls participated in the study at NIH between November 2011 and July 2019. Patients were diagnosed by at least two movement disorders specialists (including M.H.). Criteria for exclusion included comorbid neurological disease, current substance abuse, history of traumatic brain injury, active autoimmune disorder, current suicidal ideation or use of tricyclic antidepressants or antiepileptic medication. Additionally, participants could not have contraindications for MRI scanning or be pregnant. Antidepressant medication use within the past 6 months and active psychiatric disease were additional exclusion criteria for control participants. All participants provided written informed consent prior to study participation. The NIH Institutional Review Board approved the study protocol. A subset of these subjects was previously reported in Maurer et al, 2016.

### 2.2. Data Acquisition

#### 2.21. Clinical Assessment

Participants met with an experienced clinician who assessed each subject according to the Clinician Global Impression (CGI) scale. Participants completed the Beck Depression Inventory (BDI) and the State-Trait Anxiety Inventory (STAI–S).

#### 2.22. MRI Acquisition

Imaging was acquired with a 3T Siemens Skyra scanner using a 32-channel head coil. Structural images were collected using T1-weighted anatomical MRI (multi-echo magnetization-prepared rapid gradient echo [MEMPRAGE], voxel size 1 × 1 × 1 mm; repetition time [TR] 400 ms; echo time [TE] 1.69 ms; echo spacing 9.8 ms; number of echoes 4; bandwidth 650 Hz/Px; inversion time 1,100 ms; flip angle 7°; acceleration factor 2; matrix size 176 × 256 × 256; field of view [FoV] 256 mm, acquisition time: 6 minutes 2 seconds). An rs-fMRI scan was acquired during using a T2-weighted multi-echoplanar imaging sequence (voxel size 3 × 3 × 3 mm; TR 2,000 ms; TE 11/22/33 ms; flip angle 70°; FoV 210 mm, phase FoV 87.5%, acceleration factor 3, number of slices 34; interleaved, bandwidth 2,552 Hz/Px; repetitions: 180, acquisition time: 6 minutes). During these resting state scans, participants were instructed to close their eyes, remain awake, and not think of anything in particular.

### 2.3 Data Analyses

We conducted statistical testing to discover whether any differences in age, gender or clinical test scores existed between the patient and control groups. For subject age and clinical scores, a two-sample t-test was used to compare the groups. For subject gender, we used a Chi-square test.

#### 2.3.1. Image Preprocessing

Image preprocessing was completed using AFNI software (Cox, 1996). The MEMPRAGE images were averaged across echoes, then skull stripped and nonlinearly registered to the MNI ICBM 152 2009 template using the @SSwarper function. Each rs-fMRI sequence was preprocessed using afni_proc.py, where spatial transformations (*) were concatenated and applied in a single step. Preprocessing steps were as follows: despiking, slice-time correction, alignment to the processed MEMPRAGE*, registration to the MNI template space using the warps computed by @SSwarper*, co-registration of each functional volume*, multi-echo independent component analysis denoising via tedana.py (with option to not reject mid-Kappa components), convolution with a 4-mm full-width at half maximum Gaussian kernel, scaling each voxel time series to a mean of 100, and regression of 12 motion parameters (demeaned motion parameters and their first derivatives), producing residuals with a mean of zero. Volumes with excess motion (frame-wise displacement > 0.3mm) and/or with BOLD signal outliers in many voxels (proportion of voxels > 0.1) were censored. All voxels in censored timepoints were set to zero. Max displacement and number of censored TRs were extracted and inspected for differences between FMD and control groups.

#### 2.3.2 Feature Generation

66 seed locations were determined based on MNI coordinates published in prior imaging studies of FMD/FND patients (Supp. Table 2). Seed regions were spherically inflated from the central coordinate with a radius of 5 mm. In a few cases, the central coordinate was slightly adjusted away from the edge of the brain to avoid overlap with non-brain areas when inflated. We calculated the pairwise Pearson’s correlation coefficient (r) between each seed and every other seed, creating a 66 × 66 correlation matrix using AFNI software. Then, the data were inspected and reshaped using a custom Python (Python Software Foundation. Python Language Reference, version 3.7. Available at http://www.python.org) pipeline. For quality control, regions where more than 5% of subjects had zero non-null voxels were excluded from further analysis. We trimmed the full matrix to the first diagonal and removed N x N correlations, then reshaped the remaining values into a single feature vector for each subject. We inspected intra-subject feature variability (standard deviation (SD) of the feature vector) and removed any subjects whose feature variability exceeded 2.5 SD from the mean.

#### 2.3.3 Model Workflow

A two-step iterative process was deployed to reduce the number of features, then train and test the model. First, the large set of possible features was winnowed down in a feature reduction step. Second, we trained a classifier to predict the patient status of each sample. Figure 1 shows the schematic rendering of the process. First, we split the dataset into three parts: the data for feature set development and model training (80%), a validation dataset (10%) and a “hold out” dataset (10%) (figure 1). The split was performed with stratification to maintain proportionate numbers of FMD to control subjects, along with gender and age.

**Figure 1:**
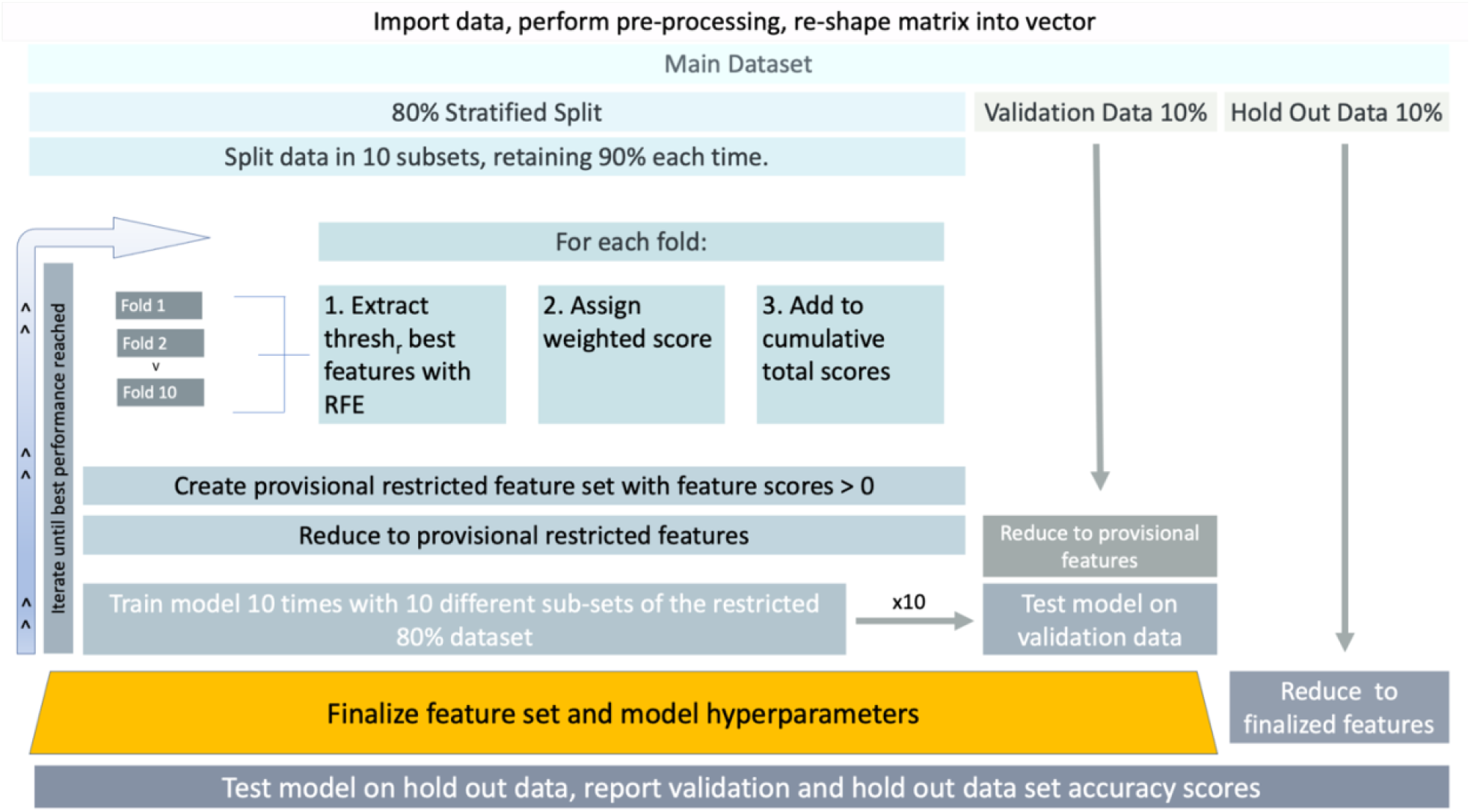
Schematic workflow for the main dataset shows the data handling steps that occurred to select a restricted feature set and train the model. <u>IN COLOR.</u>

We used an iterative recursive feature elimination (RFE) process to reduce the number of features to present during classifier model training. The RFE process uses a linear support vector machine (SVM) classifier to rank the features by order of importance (Guyon et al., 2002) and is supported by Python’s sci-kit learn package. To reduce the impact of any one sample on the selected features derived from the RFE process, we divided the 80% dataset into S sub-sets (s), where S = 10, each containing 90% of those samples. For each subset, each feature (f) was assigned a rank (f_r_) based on the RFE estimate of that feature’s importance to the model. Feature space was constrained to those below the rank threshold (thresh_r_). To preserve feature rank interpretability, these features received a feature weight (w_f_), with 1 as the maximum weight (equation 1).

Within each subset, features with f_r_ > thresh_r_ were assigned a weight of zero (equation 2). We calculated the cumulative score for each feature across the S subsets. The highest possible feature score was S (if a feature was the top ranked feature in every subset) and the maximum number of features in each restricted feature set was S x thresh_r_ (if every subset generated an unique set of ranked features above the threshold). We trimmed any feature that did not score above 0 to create a provisional restricted feature set for further evaluation.

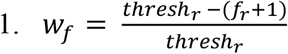

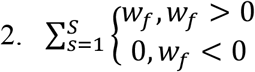

All features in the 80% dataset, along with those in the validation dataset were trimmed so that only those features identified as belonging to the provisional restricted feature set remained. We then tested model performance using a SVM with a radial basis function (rbf) by training the model on ten completely different subsets of the 80% dataset evenly balanced between FMD patients and healthy controls that were stratified to preserve gender and age ratios. We then tested model performance, measured by the performance of the model in predicting FMD versus healthy controls on the validation dataset, which contained completely independent samples that were not used during the feature selection process. The use of the validation dataset was necessary to prevent information leakage, since all samples in the 80% dataset had already been used in the RFE feature winnowing step. This process produced ten accuracy scores which were averaged to produce a single measure of model performance. Concurrently with the feature selection process, we searched for the combination of hyperparameters that best maximized the model’s performance. The composition of each provisional feature set depended on the thresh_r_ parameter in the RFE step, however the performance of individual feature sets is also determined by the hyperparameters C and gamma that determine characteristics of the rbf kernel in the final model (Vanderplas, 2016). Thus, we performed a search to determine the optimal combination of thresh_r_, C and gamma. The testing range for thresh_r_ was between two and 10, inclusive. For the C and gamma hyperparameters, we first tested with several different orders of magnitude to narrow down values which would produce an optimal solution. This yielded a search space with ranges for C of 0.1 to 2 and for gamma a range of 0.1 to 1. The thresh_r_, C and gamma values corresponding to the best performing feature set and model hyperparameters were then selected based on the empirical results of the classifier trained with the provisional restricted feature set on the validation data set. Once this optimal combination parameters and hyperparameters was determined, the contents of the restricted feature set were finalized. We reduced the number of features in the hold out dataset, which had remained completely separate during the tuning and feature selection process, to those contained in the finalized restricted feature set, then tested the performance of the model on this hold out dataset in a similar manner to the validation dataset, by observing the predictive performance of the classifier trained on the 10 subsets, then averaging the results to produce a single accuracy score.

#### 2.3.4 Feature Exploration

To highlight potential patterns in the full feature set and how these related to the restricted set of features, we performed an exploratory unsupervised K-means clustering of the full set of features (seed regions) with Python’s sci-kit learn package (Pedregosa et al., 2011) using the data from all subject samples in the main dataset. First, the optimal number of clusters were determined by generating silhouette scores for cluster numbers ranging from two to ten and selecting the number of clusters corresponding to the highest score. Second, each seed region was assigned to one of these cluster sub-groupings via the K-means clustering algorithm. Based on the sub-grouping to which each seed region was assigned, we developed a qualitative title for each sub-group using the following method: since each feature was a seed region with a central MNI coordinate, we used NeuroSynth, an automated meta-analysis software that performs text mining to match study characteristics to individual MNI coordinates (Yarkoni et al., 2011), as a source for qualitative information about each individual seed region. In a few cases, it was necessary to adjust the coordinate location by one voxel in order to generate this information. For each central coordinate, we extracted the top three descriptors (if available), excluding those that described the physical structure or brain region (such as “parietal” or “hippocampus”) or were closely associated with a particular methodology (such as “EEG”). We consolidated descriptors with highly similar meanings (such as “motion” and “movement” or “vision” and “visual”), then binned all descriptors for seed regions assigned to each cluster sub-group to better understand the relative frequency with which certain characteristics were present in each cluster. Descriptors were categorized according to similarity to generate qualitative associations.

## Results

There were no significant differences between age or gender distribution between the two groups in the main dataset. FMD patients had significantly higher CGI, BDI and STAI-S scores than healthy controls (Table 1). We removed one FMD subject from the analysis whose feature variability exceeded 2.5 SD (8.6 SD above the subject mean). There were no significant differences between the FMD and control groups in number of censored TRs, maximum displacement or average motion.

**Table 1.** Demography and clinical scores of study participants. P-value represents the results of statistical testing between the healthy control and FMD patient groups with tests described in section 2.3. Where the test was not significant, ns is indicated.

|  | FMD Patients (N = 61) | Healthy Controls (N = 59) | P-value |
| --- | --- | --- | --- |
| Mean Age (SD) | 43.8 (12.8) | 44.8 (12) | ns |
| Gender (M : F) | 12 : 49 | 14 : 45 | ns |
| Median CGI (SD) | 3 (1.35) | 1 (0.42) | > 0.001 |
| Mean BDI (SD) | 9.7 (7.7) | 2.6 (3.8) | > 0.001 |
| Mean STAI-S (SD) | 41.4 (12) | 32.9 (8.9) | > 0.001 |

We excluded three seed regions from the feature set because more than 5% of the subjects were missing data within the location. The final number of seed regions was 63. After removing the second diagonal (duplicates of the first diagonal), the total number of possible features was 1,953.

### 3.1 Feature Exploration

Unsupervised clustering of the feature set produced the highest silhouette score (0.117) with three clusters (supplementary 1). Figure 2 shows the average Pearson’s r correlation scores between each region, sorted by cluster. Visual inspection of the clusters indicates that clusters 1 and 3 were more coherent than cluster 2.

**Figure 2:**
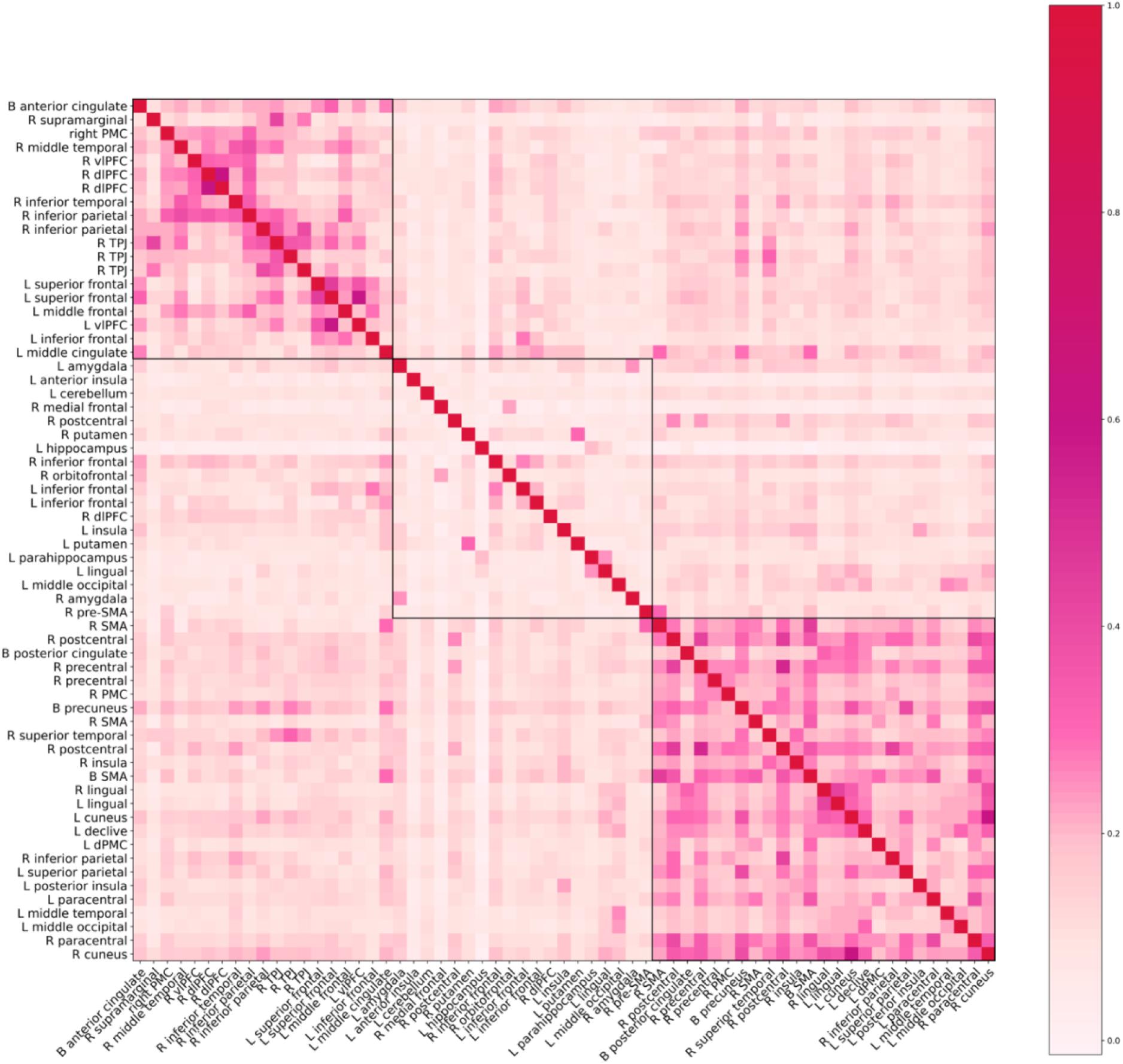
Matrix of average correlations between 63 seed regions across the full 120 sample dataset. Region order is determined by similarity to neighbors according to the unsupervised clustering algorithm. The algorithm divided the regions into three groups, which are delineated by the black borders. The lowest average r value by feature was -0.0137. All but four of the regions had positive average correlations. The maximum average r value was 0.608.

We obtained 173 total descriptors from the NeuroSynth database (some regions did not have at least three descriptors). Based on the descriptors for each ROI, we identified 19 qualitative characteristics represented within the feature set. For example, the descriptors “motor”, “movement” and “motion” were all included under the Motion/Movement characteristic. 168 of the descriptors fit within these 19 characteristic associations; 5 were excluded because the meaning was ambiguous. Table 2 shows the complete list of 19 qualitative characteristics and the frequency with which the characteristic appeared in describing the seed regions included in each group. Executive function and memory, emotion and speech, and movement were the three characteristics that were most different between groups for group 1, 2, and 3, respectively. Based on these differences, we termed the cluster groupings “Executive”, “Limbic”, and “Motor” groups.

**Table 2:** Each row represents a defined characteristic summarizing a similar grouping of descriptors, with 19 distinct characteristics. For visualization purposes, we ordered the descriptors and used a sequential color scale to show the ways the three groups differ. Numbers are the frequency with which the descriptor was present within the characteristic for each group.

|  | 1:Executive Function | 2:Limbic | 3:Motor and Motor-Related |
| --- | --- | --- | --- |
| Somatosensory/Sensorimotor |  |  | 2 |
| Motor Imagery |  |  | 6 |
| Body Part |  |  | 7 |
| Faces |  | 2 | 9 |
| Vision |  | 3 | 10 |
| Motion/Movement |  | 5 | 16 |
| Spatial Awareness/Spatial Tasks |  | 6 | 7 |
| Words, Speech, Audition | 4 | 11 | 3 |
| Affective/Emotional | 3 | 6 | 1 |
| Risk/Reward | 3 | 5 | 2 |
| Pain | 1 | 3 | 1 |
| Empathy | 4 | 2 | 2 |
| Memory | 9 | 4 |  |
| Executive Tasks | 10 | 3 |  |
| Attention | 5 |  |  |
| Thought | 4 |  |  |
| Beliefs | 3 |  |  |
| Default Mode | 2 |  | 3 |
| Social | 1 |  |  |

### 3.2 Feature Selection

Based on performance in the validation set, the optimal maximum feature threshold (thresh_r_) parameter was 3 when S = 10. 29 seed to seed connectivity features were identified for the restricted feature set (Supp. Table 3 for complete list). Of the 29 identified connections, 35 seed regions were represented. The majority of regions (31 of 35) were interconnected (more than one region to region connection) while 4 regions were isolated (one region to one region). Figure 3 shows the connectivity diagram for all seed regions. The number of regions and their interconnections were not evenly distributed within the unsupervised clustering groupings. 15 features connected a region within the motor-related cluster, 12 within the limbic-related cluster and 8 in the executive-related cluster. Seed regions that were clustered in the motor grouping (group 3) were most likely to connect either to the limbic-related cluster (group 2) or to other group 3 motor-related seeds (9 and 8 connections, respectively). Only one motor-related region connected to a region clustered in the executive function grouping (group 1). Limbic-related regions connected to other group 2 regions or group 1 regions 4 and 4 times, respectively. Executive related regions were inter-connected with other group 1 regions 3 times (Table 3).

**Table 3:**
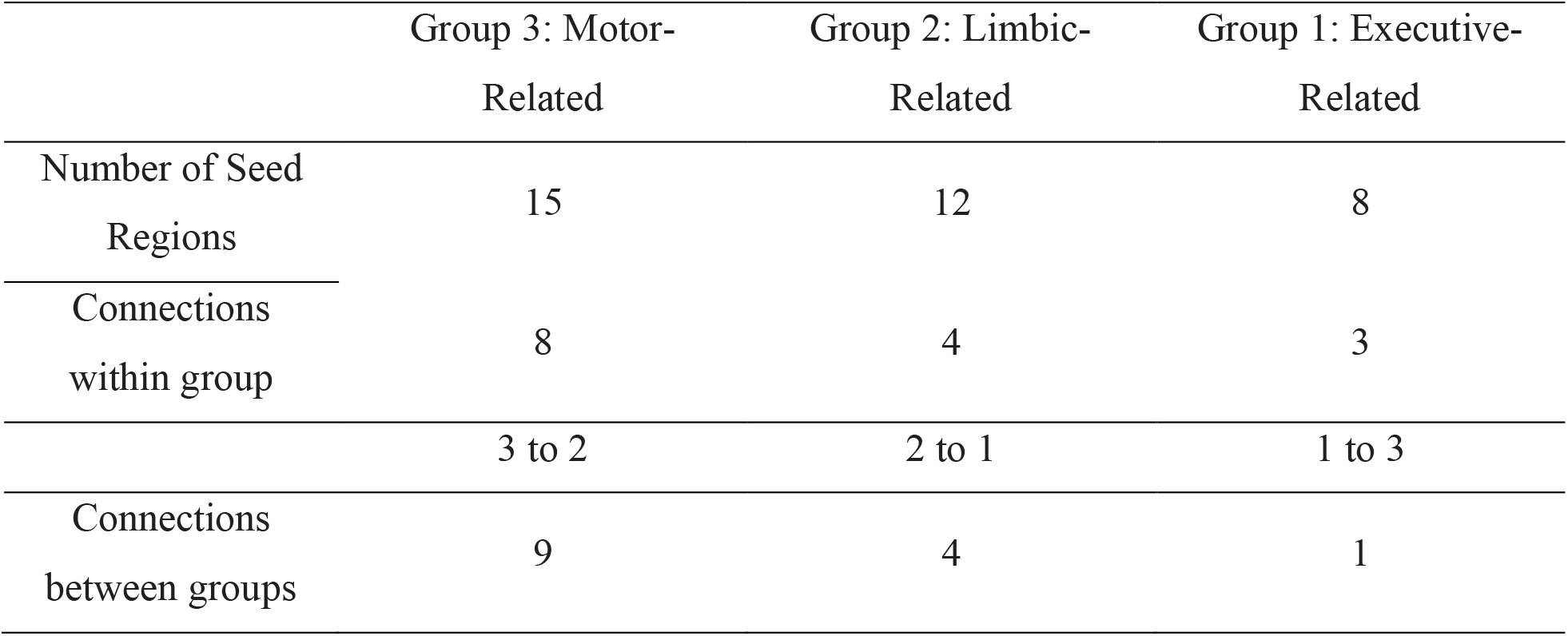
Shows the frequency of apportionment in restricted feature set between the three cluster groupings determined in the full dataset. Groups 3 and 2 had more regions represented in the restricted feature solution than group 1. Motor regions were most likely to connect either to other motor regions or group 2 regions. Limbic regions were most likely to connect to motor regions; but were equally likely to connect to other limbic and executive-related regions. Executive regions were most likely to connect to limbic regions, followed by other executive regions, with only a single direct connection to a motor region.

|  | Group 3: Motor-Related | Group 2: Limbic-Related | Group 1: Executive-Related |
| --- | --- | --- | --- |
| Number of Seed<br>Regions | 15 | 12 | 8 |
| Connections<br>within group | 8 | 4 | 3 |
|  | 3 to 2 | 2 to 1 | 1 to 3 |
| Connections<br>between groups | 9 | 4 | 1 |

**Figure 3:**
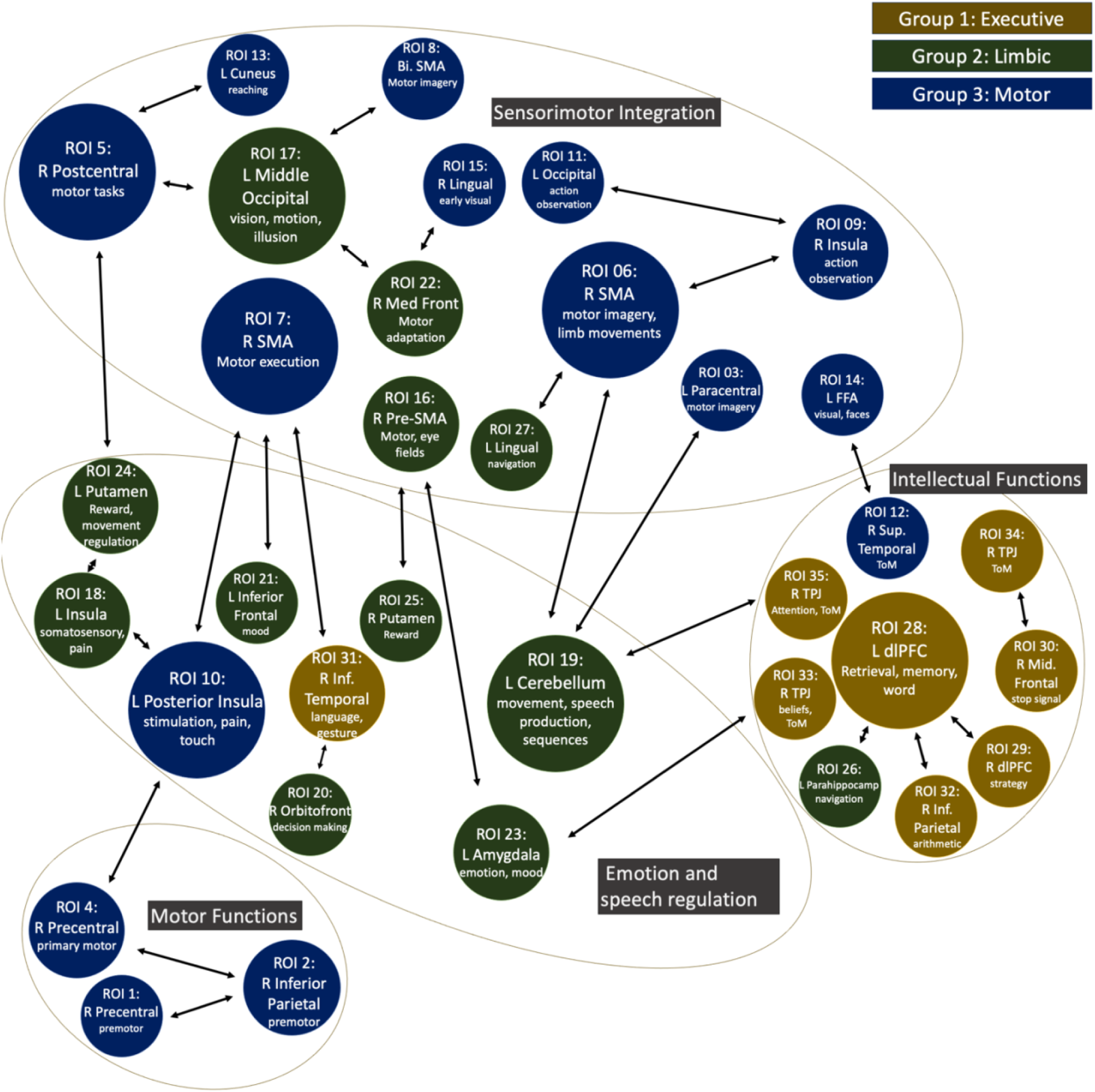
Network diagram shows the relationship between the features selected during the RFE step. Out of 1953 possible features, these 29 region-to-region connections (with 35 seed regions overall) were the best predictors of FMD patient status. For visualization purposes, regions are organized thematically according to associated functions described in NeuroSynth and their projections. Nodes are color coded according to the groupings derived from the unsupervised clustering. Node size corresponds to the number of connections attached to that node.

### 3.3 Classification Results

Average accuracy in predicting the condition of each sample in the validation dataset was 84%. When the trained model was tested on the hold out set, which had been completely held out from any model fitting or parameter/hyperparameter tuning, the classifier model had an average accuracy rate of 80% in identifying FMD subjects from healthy controls (Table 4).

**Table 4:** Accuracy rates, sensitivity and specificity percentages for the validation and hold out datasets.

|  | Validation Set (N=10) | Hold Out Set (N=11) |
| --- | --- | --- |
| Accuracy (%) | 84 | 80 |
| Sensitivity (%) | 75 | 100 |
| Specificity (%) | 100 | 60 |

## Discussion

The two-step SVM classifier was able to accurately distinguish FMD patients from healthy controls with an average accuracy rate of 80% in the hold out portion of the main dataset solely using features derived from rs-fMRI data. Through the RFE process, the model selected a limited subset of 29 region to region connections that together comprise a distinctive, if complex, pattern for predicting FMD in a broad range of patients. The dominant cluster grouping in the identified pattern was the motor-related cluster, followed by the emotion and social cognition-related (limbic) cluster. Areas associated with motor function mostly connected to other motor areas or areas associated with emotion/limbic function, with only one direct connection between a motor-related area in the right SMA and a region categorized as part of the executive function cluster in the right inferior temporal gyrus. Seed regions categorized under the limbic grouping connected more frequently both to motor regions and executive function regions. This pattern may indicate that connections between areas associated with movement, such as primary and pre-motor cortex and executive function, including areas associated with attention, theory of mind, and self-agency in the right TPJ and inhibition in the dlPFC are mostly indirect and mediated through regions associated with a series of functions, including sensorimotor integration, pain, emotion and social cognition. Interestingly, it appears that the feature selection process picked out connections between the right SMA and areas associated with a range of functions, including the left amygdala, the right putamen and the left posterior insula, which are associated with emotion and fear, emotional decision making and pain, respectively, without selecting direct connections between the SMA and pre-motor and primary motor seed regions.

The unsupervised clustering conducted on the full feature set was exploratory, however some distinct qualitative differences were evident. Groups 1 and 3 (executive and motor) appeared to have more within-group cohesion than group 2, which seemed to contain both regions connected to the limbic system, such as the amygdala, and those associated with speech and language. This may be one reason why the silhouette scores were quite low, as a less cohesive cluster can reduce the overall value. Additionally, the locations for these regions were derived from multiple studies with varying protocols, numbers of subjects and disease presentations, and this heterogeneity may have limited the underlying cohesion of the data. Nevertheless, we found these clusters helpful for providing a qualitative framework for the features and the types of functions that are implicated in FMD. Notably, these are similar in conceptualization to other researcher’s characterizations of the disorder, for example the “sensorimotor domain, the cognitive domain and the negative affect domain” (Spagnolo et al., 2021).

One problem we faced in developing the model was the number of available samples. Clinical data collection practices often collect fewer data samples than many machine learning techniques were optimized for (Arbabshirani et al., 2017). Another challenge was the large number of potential features that were initially considered. Feature winnowing required an iterative process of selecting provisional feature sets of varying compositions, then testing model performance with the validation dataset, while avoiding overfitting or excessive bias to individual samples. By employing separate validation and hold out datasets, we sought to verify the reliability of the model and avoid overfitting to small numbers.

The identification of a pattern of brain connectivity in FMD that can distinguish patients with 80% accuracy is a useful advancement toward improving diagnostic confidence. However, given the relatively small size of the dataset and the lack of external validation, this is only a first step towards developing a well-validated predictive classifier for FMD. The identified resting state markers, developed through feature winnowing, could be helpful starting point in further advancements towards a model that is robust to differences in scanner acquisition, location, and other factors that may impact classifier performance. Such an initiative might involve building a cross-site coalition to maximize the number of training samples and variation in techniques and is beyond the scope of this study.

## Conclusion

With a moderate sample size, we were able to classify FMD patients from controls with approximately 80% accuracy. The reduced group of regions defined by our feature selection, and the analysis of the relevant connections, support the concept of the FMD abnormal movement resulting from an imbalance of the limbic and motor areas.

## Data Availability

The data that support the findings of this study are available from the corresponding author, SGH, upon reasonable request and Institutional approval.

## Acknowledgements

This work was supported by the NINDS Intramural Research Program. This work utilized the computational resources of the NIH HPC Biowulf cluster (http://hpc.nih.gov) and the Functional Magnetic Resonance Imaging Facility (https://fmrif.nimh.nih.gov). Authors thank Drs. Carine Maurer and Kathrin LaFaver for participation in data acquisition and patient evaluation. Authors thank Dr. Francisco Pereira for advice and methodological guidance.

## CRediT authorship contribution statement

Rebecca Waugh: Formal analysis; Methodology; Visualization; Writing – original draft

Jacob Parker: Investigation; Writing – review & editing

Mark Hallett: Resources; Supervision; Writing – review & editing

Silvina Horovitz: Conceptualization; Investigation; Methodology; Supervision; Writing – review & editing

## Supplementary Material

**Table 1:** A feature list was derived from locations identified in a wide variety of fMRI studies of FMD/FND patients. Two studies (Maurer et al., 2016 and Baek et al., 2017) were resting state paradigms using connectivity analyses with TPJ/parietal seeds. The remainder used a range of motor and emotional tasks to look for functional differences in FMD/FND patients. In a few cases, the central seed location was slightly adjusted from the coordinates identified in the original paper to avoid intersecting with edge of the brain (demarked with an asterix (*)). Coordinates from the Espay et al. (2018) paper are approximations based on published images (demarked with ^**I**^).

| Label | Seed Region | MNI Coordinates | Source |
| --- | --- | --- | --- |
| ROI 1 | R Precentral | 38, -12, 39 | Espay et al., 2018 <sup>i</sup> |
| ROI 2 | R Inferior Parietal | 36, -38, 43 | Maurer et al., 2016 |
| ROI 3 | L Paracentral | -6, -20, 70 | Stone et al., 2007 |
| ROI 4 | R Precentral | 48, -17, 55 | Maurer et al., 2016 |
| ROI 5 | R Postcentral | 56, -23, 52 | Maurer et al., 2016 |
| ROI 6 | R SMA | 12, -8, 68 | Aybek et al., 2014 |
| ROI 7 | R SMA | 6, 3, 56 | Baek et al., 2017 |
| ROI 8 | B SMA | 0, -8, 61 | Maurer et al., 2016 |
| ROI 9 | R Insula | 56, -34, 19 | Maurer et al., 2016 |
| ROI 10 | L Posterior Insula | -38, -24, 20 | Stone et al., 2007 |
| ROI 11 | L Middle Temporal | -45, -65, 5 | Maurer et al., 2016 |
| ROI 12 | R Superior Temporal | 52, -44, 20 | Aybek et al., 2014 |
| ROI 13 | L Cuneus | -15, -80, 40 | Baek et al., 2017 |
| ROI 14 | L Middle Occipital | -42, -77, -11 | Maurer et al., 2016 <sup>*</sup> |
| ROI 15 | R Lingual | 10, -68, 2 | Stone et al., 2007 |
| ROI 16 | R SMA | 10, 6, 60 | Voon et al., 2010 <sup>*</sup> |
| ROI 17 | L Middle Occipital | -40, -80, 2 | Baek et al., 2017 <sup>*</sup> |
| ROI 18 | L Insula | -46, -18, 10 | Espay et al., 2018 <sup>i</sup> |
| ROI 19 | L Cerebellum | -13, -69, 10 | Baek et al., 2017 |
| ROI 20 | R Orbitofrontal | 12, 50, -6 | Stone et al., 2007 |
| ROI 21 | L Inferior Frontal | -40, 26, -8 | Stone et al., 2007 |
| ROI 22 | R Medial Frontal | 20, 45 -3 | Baek et al., 2017 |
| ROI 23 | L Amygdala | -23, -5, -15 | multiple |
| ROI 24 | L Putamen | -26, 14, -2 | Stone et al., 2007 |
| ROI 25 | R Putamen | 20, 12, -4 | Stone et al., 2007 |
| ROI 26 | L Parahippocampus | -22, -44, 0 | Aybek et al., 2014 |
| ROI 27 | L Lingual | -26, -46, -4 | Aybek et al., 2014 |
| ROI 28 | L Middle Frontal | -52, 19, 34 | Maurer et al., 2016 * |
| ROI 29 | R Middle Frontal | 48, 46, 18 | Stone et al., 2007 |
| ROI 30 | R Middle Frontal | 38, 53, 2 | Baek et al., 2017 |
| ROI 31 | R Inferior Temporal | 60, -55, -14 | Baek et al., 2017 * |
| ROI 32 | R Inferior Parietal | 41, -65, 58 | Baek et al., 2017 |
| ROI 33 | R Inferior Parietal | 48, -55, 32 | Baek et al., 2017 |
| ROI 34 | R TPJ | 51, -46, 31 | Maurer et al., 2016 |
| ROI 35 | R Angular | 40, -58, -24 | Aybek et al., 2014 |
| ROI 36 | B Posterior Cingulate | 0, -64, 7 | Maurer et al., 2016 * |
| ROI 37 | B Precuneus | 0, -52, 58 | Maurer et al., 2016 * |
| ROI 38 | L Middle Cingulate | -3, 19, 37 | Maurer et al., 2016 |
| ROI 39 | L Declive | -36, -65, -19 | Maurer et al., 2016 |
| ROI 40 | R Cerebellum | 31, -65, -54 | Baek et al., 2017 |
| ROI 41 | R Amygdala | 23, -5, -15 | multiple |
| ROI 42 | R Supramarginal | 42, -54, 30 | Aybek et al., 2014 |
| ROI 43 | L Hippocampus | -28, -42, 2 | Aybek et al., 2014 |
| ROI 44 | B Anterior Cingulate | 1, 45, 10 | Espay et al., 2018 <sup>i</sup> |
| ROI 45 | L Anterior Insula | -24, 20, 14 | Stone et al., 2007 |
| ROI 46 | R Middle Temporal | 68, -34, -10 | Maurer et al., 2016 |
| ROI 47 | L Superior Frontal | 0, 57, 31 | Maurer et al., 2016 |
| ROI 48 | L Superior Frontal | -15, 55, 31 | Maurer et al., 2016 * |
| ROI 49 | R Cuneus | 15, -80, 40 | Baek et al., 2017 * |
| ROI 50 | R Premotor | 55, 0, 35 | Baek et al., 2017 |
| ROI 51 | R Middle Temporal | 62, -25, -19 | Baek et al., 2017 |
| ROI 52 | R Middle Frontal | 31, 28, 35 | Baek et al., 2017 |
| ROI 52 | R Inferior Parietal | 43, -69, 30 | Baek et al., 2017 |
| ROI 53 | R Premotor | 36, 7, 58 | Baek et al., 2017 * |
| ROI 54 | L Cerebellum | -48, -55, 32 | Baek et al., 2017 |
| ROI 55 | R Postcentral | 30, -42, 65 | Aybek et al., 2014 * |
| ROI 56 | R Postcentral | 28, -34, 54 | Aybek et al., 2014 |
| ROI 57 | R Prefrontal | 6, 56, 32 | Dognowski et al., 2019 * |
| ROI 58 | L Inferior Frontal | -46, 38, -6 | Stone et al. , 2007 |
| ROI 59 | L Inferior Frontal | -44, 16, 6 | Stone et al., 2007 |
| ROI 60 | L Lingual | -12, -78, 2 | Stone et al., 2007 |
| ROI 61 | R Inferior Frontal | 40, 20, -10 | Stone et al., 2007 |
| ROI 62 | L Superior Parietal | -14, -60, 62 | Stone et al., 2007 |
| ROI 63 | R Paracentral | 8, -44, 68 | Stone et al., 2007 |
| ROI 64 | R Middle Frontal | 38, 58, 2 | Stone et al., 2007 * |
| ROI 65 | R Premotor | 14, -4, 70 | Dognowski et al., 2019 * |
| ROI 66 | L Premotor | -20, -8, 68 | Dognowski et al., 2019 * |

**Table 2:**
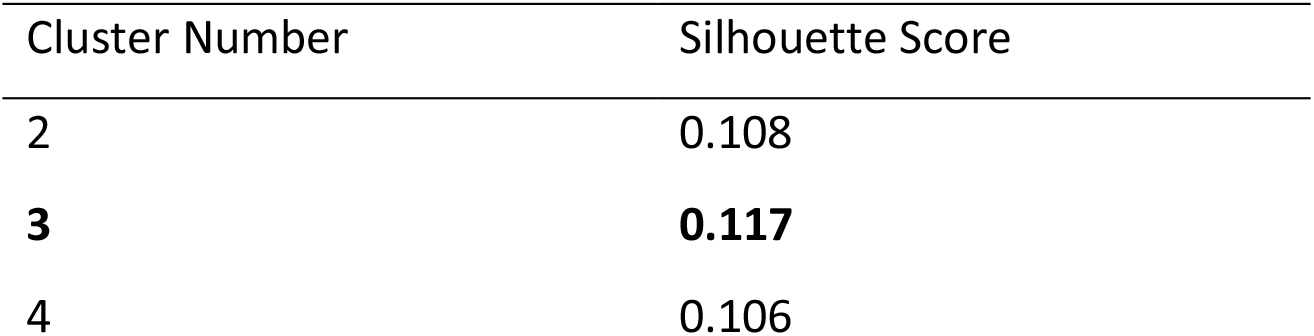
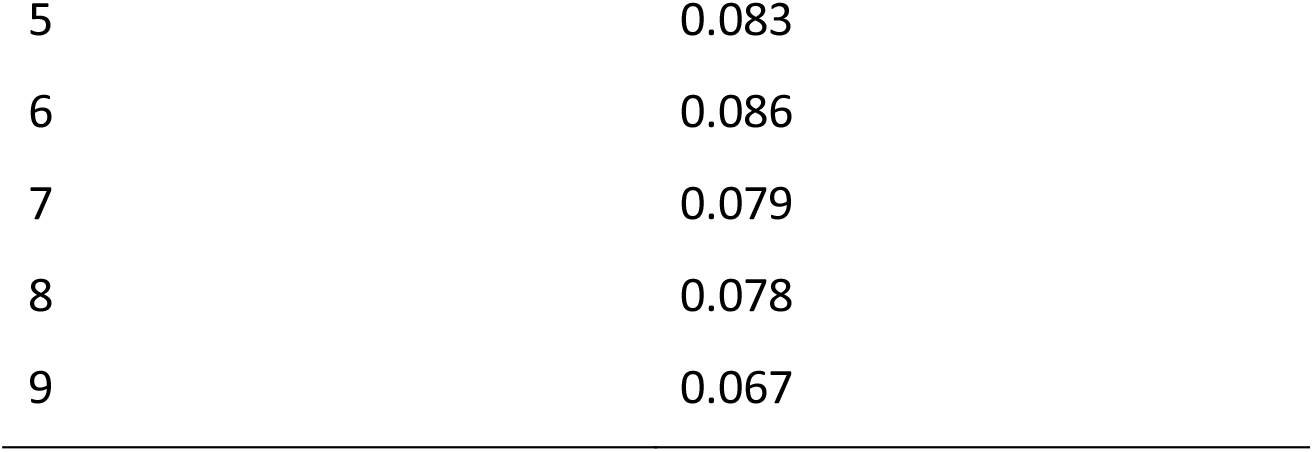
Silhouette scores measure the coherence of individual clusters in unsupervised K-Means clustering of the full feature set and aid in specifying the appropriate number of clusters. The highest silhouette score (in bold) was with three clusters.

| Cluster Number | Silhouette Score |
| --- | --- |
| 2 | 0.108 |
| **3** | **0.117** |
| 4 | 0.106 |
| 5 | 0.083 |
| 6 | 0.086 |
| 7 | 0.079 |
| 8 | 0.078 |
| 9 | 0.067 |

**Table 3:** Complete list of 29 ROI to ROI connections selected during RFE feature reduction process. Weighted scores are the cumulative scores across all 10 subsets that were tested.

| Seed Region 1 | Seed Region 2 | Weighted Score |
| --- | --- | --- |
| ROI 04: R Precentral | ROI 10: L Posterior Insula | 1.667 |
| ROI 02: R Inferior Parietal | ROI 01: R Precentral | 1 |
| ROI 05: R Postcentral | ROI 13: L Cuneus | 1 |
| ROI 28: L Middle Frontal | ROI 32: R Inferior Parietal | 1 |
| ROI 19: L Cerebellum | ROI 06: R SMA | 1 |
| ROI 17: L Middle Occipital | ROI 22: R Medial Frontal | 1 |
| ROI 31: R Inferior Temporal | ROI 20: R Orbitofrontal | 1 |
| ROI 23: L Amygdala | ROI 16: R SMA | 1 |
| ROI 16: R SMA | ROI 24: R Putamen | 1 |
| ROI 06: R SMA | ROI 27: L Lingual | 1 |
| ROI 05: R Postcentral | ROI 17: L Middle Occipital | 0.667 |
| ROI 28: L Middle Frontal | ROI 29: R Middle Frontal | 0.667 |
| ROI 34: R TPJ | ROI 30: R Middle Frontal | 0.667 |
| ROI 19: L Cerebellum | ROI 03: L Paracentral | 0.667 |
| ROI 07: R SMA | ROI 31: R Inferior Temporal | 0.667 |
| ROI 33: R Inferior Parietal | ROI 23: L Amygdala | 0.667 |
| ROI22: R Medial Frontal | ROI 15: R Lingual | 0.667 |
| ROI 18: L Insula | ROI 10: L Posterior Insula | 0.667 |
| ROI 18: L Insula | ROI 24: L Putamen | 0.667 |
| ROI 11: L Middle Temporal | ROI 09: R Insula | 0.333 |
| ROI 09: R Insula | ROI 06: R SMA | 0.333 |
| ROI 02: R Inferior Parietal | ROI 04: R Precentral | 0.333 |
| ROI 08: Bilateral SMA | ROI 17: L Middle Occipital | 0.333 |
| ROI 14: L Middle Occipital | ROI 12: R Superior Temporal | 0.333 |
| ROI 05: R Postcentral | ROI 24: L Putamen | 0.333 |
| ROI 28: L Middle Frontal | ROI 26: L Parahippocampus | 0.333 |
| ROI 19: L Cerebellum | ROI 35: R Angular | 0.333 |
| ROI 07: R SMA | ROI 10: L Posterior Insula | 0.333 |
| ROI 07: R SMA | ROI 21: L Inferior Frontal | 0.333 |

## Notes

### Competing Interest Statement

The authors have declared no competing interest.

### Author Declarations

The NIH Institutional Review Board approved the study protocol.

## References

American Psychiatric Association. (2013). Somatic Symptom and Related Disorders. In Diagnostic and Statistical Manual of Mental Disorders. https://doi.org/10.1176/appi.books.9780890425596.dsm09

Arbabshirani, M. R., Plis, S., Sui, J., & Calhoun, V. D. (2017). Single subject prediction of brain disorders in neuroimaging: Promises and pitfalls. Neuroimage, 145(Pt B), 137–165. https://doi.org/10.1016/j.neuroimage.2016.02.079

Aybek, S., Nicholson, T. R., O’Daly, O., Zelaya, F., Kanaan, R. A., & David, A. S. (2015). Emotion-motion interactions in conversion disorder: an FMRI study. PLoS One, 10(4), e0123273. https://doi.org/10.1371/journal.pone.0123273

Aybek, S., Nicholson, T. R., Zelaya, F., O’Daly, O. G., Craig, T. J., David, A. S., & Kanaan, R. A. (2014). Neural correlates of recall of life events in conversion disorder. JAMA Psychiatry, 71(1), 52–60. https://doi.org/10.1001/jamapsychiatry.2013.2842

Baek, K., Donamayor, N., Morris, L. S., Strelchuk, D., Mitchell, S., Mikheenko, Y., Yeoh, S. Y., Phillips, W., Zandi, M., Jenaway, A., Walsh, C., & Voon, V. (2017). Impaired awareness of motor intention in functional neurological disorder: implications for voluntary and functional movement. Psychol Med, 47(9), 1624–1636. https://doi.org/10.1017/S0033291717000071

Burgmer, M., Konrad, C., Jansen, A., Kugel, H., Sommer, J., Heindel, W., Ringelstein, E. B., Heuft, G., & Knecht, S. (2006). Abnormal brain activation during movement observation in patients with conversion paralysis. Neuroimage, 29(4), 1336–1343. https://doi.org/10.1016/j.neuroimage.2005.08.033

Cojan, Y., Waber, L., Carruzzo, A., & Vuilleumier, P. (2009). Motor inhibition in hysterical conversion paralysis. Neuroimage, 47(3), 1026–1037. https://doi.org/10.1016/j.neuroimage.2009.05.023

Cox, R. (1996). AFNI: Software for Analysis and Visualization of Functional Magnetic Resonance Neuroimages. Computers and Biomedical Research, 29, 162–173.

Czarnecki, K., & Hallett, M. (2012). Functional (psychogenic) movement disorders. Curr Opin Neurol, 25(4), 507–512. https://doi.org/10.1097/WCO.0b013e3283551bc1

Diez, I., Ortiz-Teran, L., Williams, B., Jalilianhasanpour, R., Ospina, J. P., Dickerson, B. C., Keshavan, M. S., LaFrance, W. C., Jr., Sepulcre, J., & Perez, D. L. (2019). Corticolimbic fast-tracking: enhanced multimodal integration in functional neurological disorder. J Neurol Neurosurg Psychiatry, 90(8), 929–938. https://doi.org/10.1136/jnnp-2018-319657

Dogonowski, A. M., Andersen, K. W., Sellebjerg, F., Schreiber, K., Madsen, K. H., & Siebner, H. R. (2019). Functional neuroimaging of recovery from motor conversion disorder: A case report. Neuroimage, 190, 269–274. https://doi.org/10.1016/j.neuroimage.2018.03.061

Espay, A. J., Maloney, T., Vannest, J., Norris, M. M., Eliassen, J. C., Neefus, E., Allendorfer, J. B., Chen, R., & Szaflarski, J. P. (2018). Dysfunction in emotion processing underlies functional (psychogenic) dystonia. Mov Disord, 33(1), 136–145. https://doi.org/10.1002/mds.27217

Faul, L., Knight, L. K., Espay, A. J., Depue, B. E., & LaFaver, K. (2020). Neural activity in functional movement disorders after inpatient rehabilitation. Psychiatry Res Neuroimaging, 303, 111125. https://doi.org/10.1016/j.pscychresns.2020.111125

Guyon, I., Weston, J., & Barnhill, S. (2002). Gene selection for cancer classification using support vector machines. Machine Learning, 46, 389–422.

Hassa, T., Sebastian, A., Liepert, J., Weiller, C., Schmidt, R., & Tuscher, O. (2017). Symptom-specific amygdala hyperactivity modulates motor control network in conversion disorder. Neuroimage Clin, 15, 143–150. https://doi.org/10.1016/j.nicl.2017.04.004

Kanaan, R. A., Craig, T. K., Wessely, S. C., & David, A. S. (2007). Imaging repressed memories in motor conversion disorder. Psychosom Med, 69(2), 202–205. https://doi.org/10.1097/PSY.0b013e31802e4297

LaFaver, K., Lang, A. E., Stone, J., Morgante, F., Edwards, M., Lidstone, S., Maurer, C. W., Hallett, M., Dwivedi, A. K., & Espay, A. J. (2020). Opinions and clinical practices related to diagnosing and managing functional (psychogenic) movement disorders: changes in the last decade. Eur J Neurol, 27(6), 975–984. https://doi.org/10.1111/ene.14200

Marshall, J. C., Halligan, P. W., Fink, G. R., Wade, D. T., & Frackowiak, R. S. J. (1997). The functional anatomy of a hysterical paralysis. Cognition, 64(1), B1–B8. https://doi.org/10.1016/S0010-0277(97)00020-6

Maurer, C., LaFaver, K., Ameli, R., Epstein, S., Hallett, M., & Horowitz, S. (2016). Impaired Self-Agency in Functional Movement Disorders. Neurology, 87, 564–570. https://doi.org/DOI10.1212/WNL.0000000000002940

Monsa, R., Peer, M., & Arzy, S. (2018). Self-reference, emotion inhibition and somatosensory disturbance: preliminary investigation of network perturbations in conversion disorder. Eur J Neurol, 25(6), 888–e862. https://doi.org/10.1111/ene.13613

Nahab, F. B., Kundu, P., Gallea, C., Kakareka, J., Pursley, R., Pohida, T., Miletta, N., Friedman, J., & Hallett, M. (2011). The neural processes underlying self-agency. Cereb Cortex, 21(1), 48–55. https://doi.org/10.1093/cercor/bhq059

Nahab, F. B., Kundu, P., Maurer, C., Shen, Q., & Hallett, M. (2017). Impaired sense of agency in functional movement disorders: An fMRI study. PLoS One, 12(4), e0172502. https://doi.org/10.1371/journal.pone.0172502

Pedregosa, F., Varoquaux, G., Gramfort, A., Michel, A., & Thirion, B. (2011). Scitkit-learn: Machine Learning in Python. Journal of Machine Learning Research, 12, 2825–2830.

Perez, D. L., Aybek, S., Popkirov, S., Kozlowska, K., Stephen, C. D., Anderson, J., Shura, R., Ducharme, S., Carson, A., Hallett, M., Nicholson, T. R., Stone, J., LaFrance, W. C., Jr., & Voon, V. (2021). A Review and Expert Opinion on the Neuropsychiatric Assessment of Motor Functional Neurological Disorders. J Neuropsychiatry Clin Neurosci, 33(1), 14–26. https://doi.org/10.1176/appi.neuropsych.19120357

Roelofs, J. J., Teodoro, T., & Edwards, M. J. (2019). Neuroimaging in Functional Movement Disorders. Curr Neurol Neurosci Rep, 19(3), 12. https://doi.org/10.1007/s11910-019-0926-y

Schrag, A. E., Mehta, A. R., Bhatia, K. P., Brown, R. J., Frackowiak, R. S., Trimble, M. R., Ward, N. S., & Rowe, J. B. (2013). The functional neuroimaging correlates of psychogenic versus organic dystonia. Brain, 136(Pt 3), 770-781. https://doi.org/10.1093/brain/awt008

Spagnolo, P. A., Garvey, M., & Hallett, M. (2021). A dimensional approach to functional movement disorders: Heresy or opportunity. Neuroscience & Biobehavioral Reviews, 127, 25–36. https://doi.org/10.1016/j.neubiorev.2021.04.005

Spence, S. A., Crimlisk, H. L., Cope, H., Ron, M. A., & Grasby, P. M. (2000). Discrete neurophysiological correlates in prefrontal cortex during hysterical and feigned disorder of movement. The Lancet, 355(9211), 1243–1244. https://doi.org/10.1016/s0140-6736(00)02096-1

Stone, J., Zeman, A., Simonotto, E., Meyer, M., Azuma, R., Flett, S., & Sharpe, M. (2007). FMRI in patients with motor conversion symptoms and controls with simulated weakness. Psychosom Med, 69(9), 961–969. https://doi.org/10.1097/PSY.0b013e31815b6c14

van Beilen, M., de Jong, B. M., Gieteling, E. W., Renken, R., & Leenders, K. L. (2011). Abnormal parietal function in conversion paresis. PLoS One, 6(10), e25918. https://doi.org/10.1371/journal.pone.0025918

Vanderplas, J. T. (2016). Python Data Science Handbook: essential tools for working with data (1 ed.). O’Reilly Media.

Voon, V., Brezing, C., Gallea, C., Ameli, R., Roelofs, K., LaFrance, W. C., Jr., & Hallett, M. (2010). Emotional stimuli and motor conversion disorder. Brain, 133(Pt 5), 1526–1536. https://doi.org/10.1093/brain/awq054

Voon, V., Brezing, C., Gallea, C., & Hallett, M. (2011). Aberrant supplementary motor complex and limbic activity during motor preparation in motor conversion disorder. Mov Disord, 26(13), 2396–2403. https://doi.org/10.1002/mds.23890

Voon, V., Cavanna, A. E., Coburn, K., Sampson, S., Reeve, A., & LaFrance, W. C., Jr. (2016). Functional Neuroanatomy and Neurophysiology of Functional Neurological Disorders (Conversion Disorder). J Neuropsychiatry Clin Neurosci, 28(3), 168–190. https://doi.org/10.1176/appi.neuropsych.14090217

Voon, V., Gallea, C., Hattori, N., Bruno, M., Ekanayake, V., & Hallett, M. (2010). The Involuntary Nature of Conversion Disorder. Neurology, 74, 223–228.

Wegrzyk, J., Kebets, V., Richiardi, J., Galli, S., de Ville, D. V., & Aybek, S. (2018). Identifying motor functional neurological disorder using resting-state functional connectivity. Neuroimage Clin, 17, 163–168. https://doi.org/10.1016/j.nicl.2017.10.012

Yarkoni, T., Poldrack, R. A., Nichols, T. E., Van Essen, D. C., & Wager, T. D. (2011). Large-scale automated synthesis of human functional neuroimaging data. Nat Methods, 8(8), 665–670. https://doi.org/10.1038/nmeth.1635

